# Implementing an Improved STEMI Alert Notification System Workflow to Reduce Alarm Fatigue in a Community Hospital

**DOI:** 10.1101/2025.05.27.25328465

**Authors:** Jomel Patrick Jacinto, Sadhana Jonna, Rayan Y Alsaghir, Syeda H Hassan, Mubbasher A Syed

**Author notes:** Corresponding author: Mubbasher A Syed, MD, MPH, FACC, FSCAI, Department of Interventional Cardiology, HCA Orange Park Hospital.

## Abstract

**Background:** The assumed national average of STEMI cancellations due to inappropriate activations is approximately 15%. In the final quarter of 2022, 57.9% of hospital-wide and 55.6% of ED/EMS STEMI activations were canceled at HCA Florida Orange Park Hospital. This cancellation rate contributed to overall “alarm fatigue” and hospital staff dissatisfaction over the hospital’s overuse of hospital resources.

**Methods:** An evaluation of the STEMI activation system revealed breakdown in communication from EMS to ED physician, inaccurate cardiology on-call schedule, and inefficient communication of case details between EMS, ED physician, and cardiologist.

An algorithm was created that directed all STEMI activations from EMS to the ED physician for review, who may activate the STEMI alert or discuss the case with the on-call cardiologist. This plan was implemented and allowed a 30-day adjustment period. After this period, a prospective analysis of STEMI cancellations in the 2nd quarter of 2023 was performed with cath lab staff satisfaction surveys.

**Results:** The data on retrospective analysis of the final quarter of 2022 showed a STEMI cancellation rate of 68.8% hospital-wide and 66.7% from the ED and EMS. After the 30-day implementation of the plan, the hospital-wide STEMI cancellation rate dropped to 20% and 22% from the ED and EMS in the 2nd quarter of 2023 without adverse effect on door-to-balloon time.

**Conclusions:** The multidisciplinary approach to the creation and implementation of the new STEMI activation algorithm resulted in a significant reduction in false activations, improved cath lab staff satisfaction, and better utilization of hospital resources.

## Introduction

The standard goal for door-to-balloon time is less than 90 minutes from the time a STEMI patient presents to an ED.(1)In the U.S., it has been observed that most adults (75%) reside near a hospital that does not offer PCI services. (2) According to recent 2025 ACC/AHA guidelines for ACS, best clinical practice includes patients calling emergency services, EMS obtaining prehospital ECG within 10 minutes of first medical contact, activating cardiac catheterization laboratory from field for suspected STEMIs, transporting patient to nearest PCI center, all from FMC to door time of 90 minutes. Communities involved in STEMI care should continue to assess and maintain regional systems to improve outcomes for patients. Key strategies for successful STEMI systems include public education for activating EMS, data collection, efficient data sharing across hospitals and EMS, and regular interdisciplinary meetings to review shared data. (3)

While EKG interpretation via cardiac monitor at the scene is quick, false positives to a PCI center remain high. (4) Additionally, early notification and prehospital CCL activation can streamline treatment, reduce delays, and alleviate logistical challenges, particularly during periods of limited staffing, such as nights and weekends. However, a commonly acknowledged limitation of prehospital CCL activation is the frequent cancellation of alerts after EMS has initiated them from the field. Contributing factors include systemic factors, including misinterpretation of the ECG, over-reliance on computer-generated readings, and challenges associated with the transmission of prehospital ECGs.

A false positive STEMI alert can cause many issues, including the risk of motor vehicle accidents, utilization of personnel and resources at the hospital, and fewer available units due to unnecessary occupancy and alarm fatigue among healthcare providers. (5)In the final quarter of 2022, at HCAFL-OPH, 57.9% of hospital-wide and 55.6% ED/EMS STEMI activations were canceled. This contributed to overall “alarm fatigue” and dissatisfaction of hospital staff, as well as poor usage of hospital resources. Reducing the number of false-positive STEMI triages in the field could lead to more efficient use of resources and enhance the safety of EMS operations.

Our objective is to create a more efficient and streamlined STEMI activation algorithm and assess pre- and post-implementation satisfaction scores among healthcare staff.

## Methodology

We conducted an analysis of all STEMI alert cancellations during the fourth quarter of 2022 to identify the underlying causes of inappropriate activation. This investigation revealed several factors contributing to false activations, including re-interpretation of EKGs by physicians and patients being poor candidates for cardiac catheterization. A comprehensive evaluation of the existing STEMI activation process identified significant communication breakdowns between EMS and the ED physician, discrepancies in the cardiology on-call schedule, and inefficiencies in the transmission of case details between EMS, the ED physician, and the on-call cardiologist.

In response to these findings, a strategic plan was developed to establish a more efficient and streamlined approach to STEMI activations. An algorithm (Figure 1), created and reviewed by all members, was submitted to the hospital’s quality department and subsequently approved for implementation. This algorithm outlined a revised protocol wherein all STEMI activations from EMS or the ED would be directed to the ED physician for review. The ED physician would then have the option to either activate the STEMI alert or consult with the on-call interventional cardiologist. To further reduce inconsistencies, the management of the cardiology on-call schedule was transferred to the physician practice, thereby improving communication and minimizing scheduling errors.

**Figure 1:**
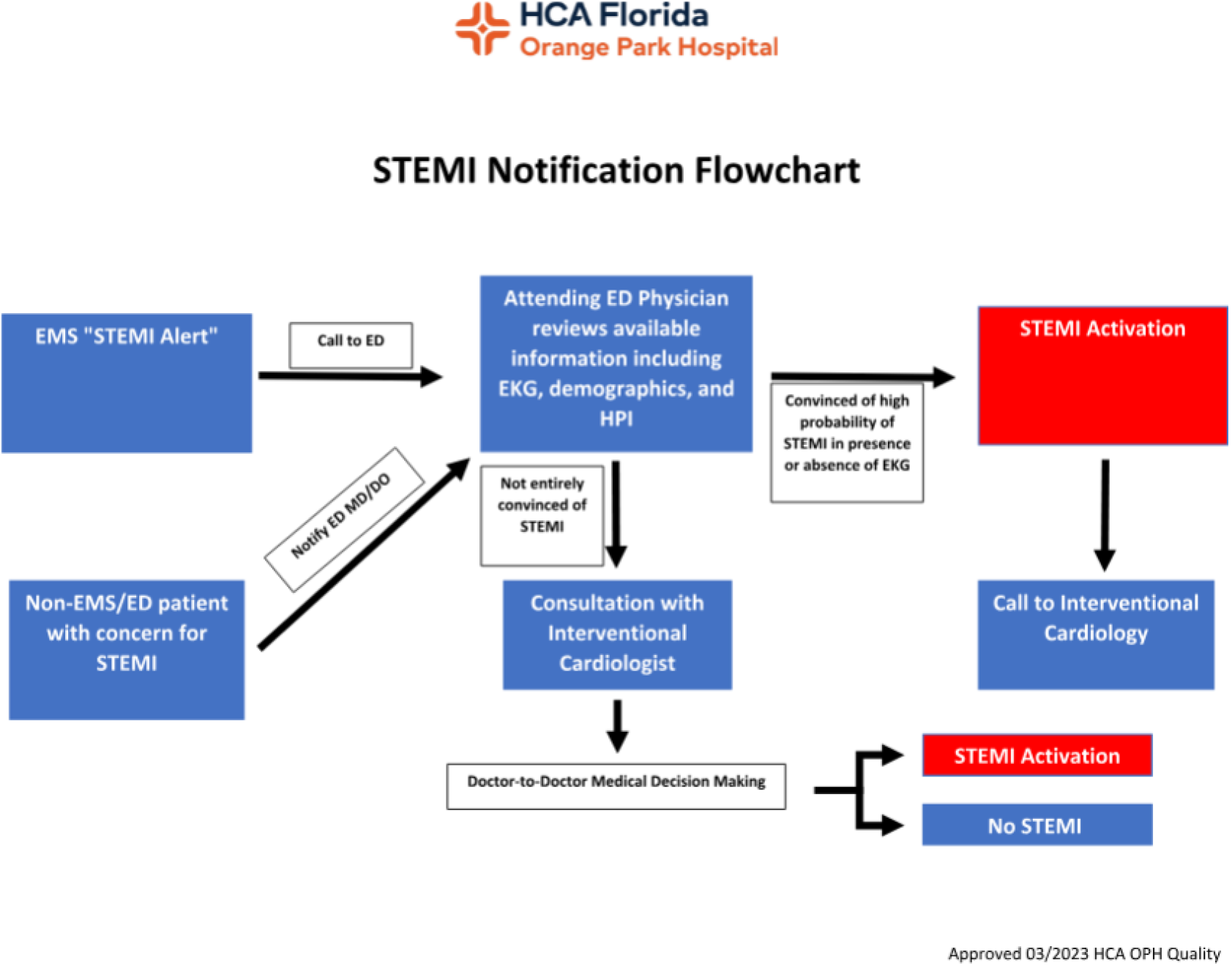
STEMI notification flowchart

Following the implementation of these changes, a 30-day adjustment period was allowed. Subsequently, a prospective analysis of STEMI cancellations during the second quarter of 2023 was conducted. Additionally, to evaluate the overall satisfaction of Cath lab staff with the new process, a seven-question survey was distributed to the staff prior to the implementation of the changes. The same survey was conducted again three months after the changes were enacted to assess any differences in satisfaction and perceptions of the new protocol.

## Results

The data on retrospective analysis of the final quarter of 2022 showed a STEMI cancellation rate of 68.8% hospital-wide and 66.7% from the ED and EMS. After the 30-day implementation of the plan, the hospital-wide STEMI cancellation rate dropped to 20% and 22% from the ED and EMS in the 2nd quarter of 2023(Tables, Table 2). Additional observations included an increase in STEMI activations from the ED and fewer activations from EMS. Furthermore, we found that the overall number of STEMI alerts activated by EMS dropped from 22 to 6 (72.7%).

**Table 1.** Pre-Intervention 4th Quarter 2022 (October - December) CCL Data:

| STEMI Location | Total CCL Activation | Total STEMI | CCL Activation (other than STEMI) | Number of Canceled STEMI | STEMI Cancellation Rate |
| --- | --- | --- | --- | --- | --- |
| EMS/Field | 22 | 20 | 2 | 11 | 55% |
| Emergency Department | 14 | 10 | 4 | 9 | 90% |
| Inpatient Floors | 2 | 2 | 0 | 2 | 100% |
| Overall Total | 38 | 32 | 6 | 22 | 68.8% (Hospital-Wide) |

**Table 2.**
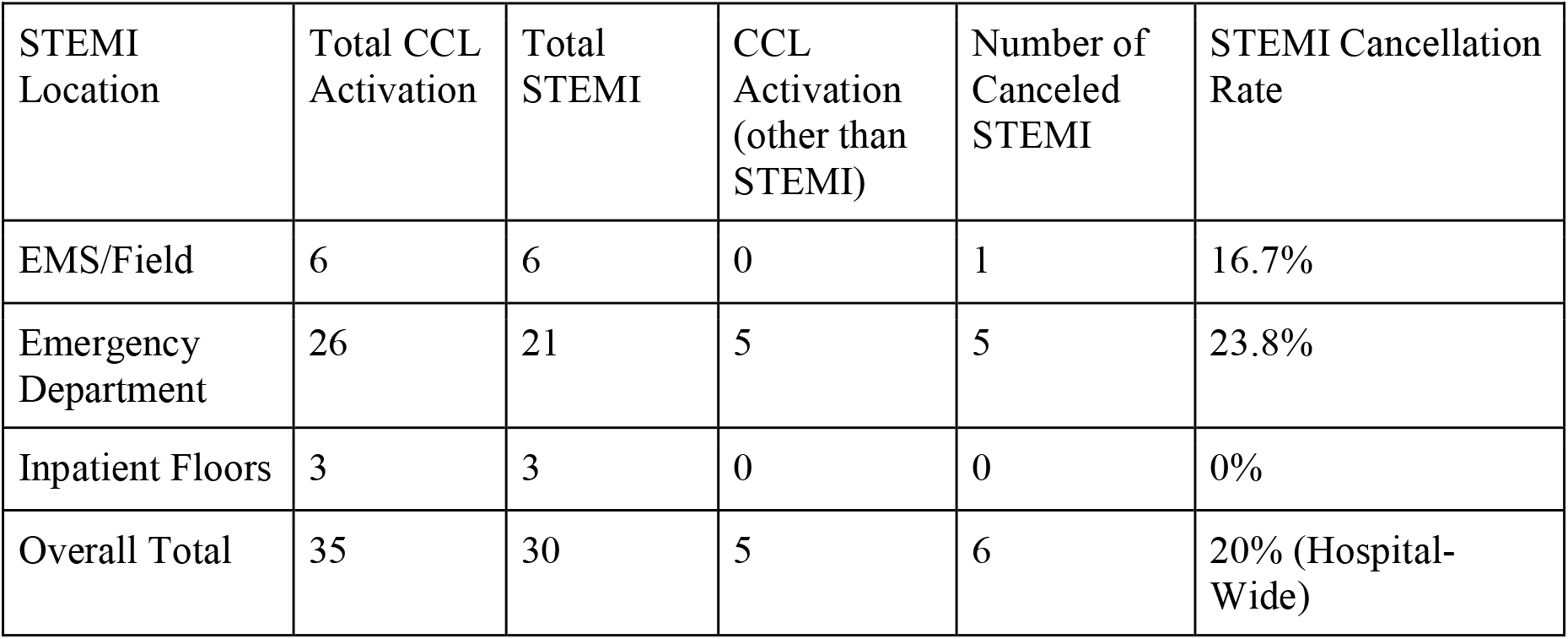
Post Intervention 2nd Quarter 2023 (April - June) CCL Data.

| STEMI Location | Total CCL Activation | Total STEMI | CCL Activation (other than STEMI) | Number of Canceled STEMI | STEMI Cancellation Rate |
| --- | --- | --- | --- | --- | --- |
| EMS/Field | 6 | 6 | 0 | 1 | 16.7% |
| Emergency Department | 26 | 21 | 5 | 5 | 23.8% |
| Inpatient Floors | 3 | 3 | 0 | 0 | 0% |
| Overall Total | 35 | 30 | 5 | 6 | 20% (Hospital-Wide) |

The primary reasons for cancellation fell into two main categories, including re-interpretation of EKG by the ED physician or cardiologist and poor patient CCL candidacy. Of the 22 cancellations of the 2022 final quarter, 18 were canceled due to reinterpretation, and 4fourwere canceled due to poor patient CCL candidacy. Of the six cancellations in the second quarter of 2023, 5 were canceled due to reinterpretation, and one was canceled due to poor patient CCL candidacy.

**Table 3.** Door-to-balloon Times.

|  | 4th Quarter 2022 | 2nd Quarter 2023 |
| --- | --- | --- |
| Median Time (minutes) | 48.5 | 68.0 |
| Average Time (minutes) | 47.13 | 66.31 |

\*Analysis of cases revealed longer D2B times due to unforeseen delays unrelated to the implemented algorithm, such as vascular access delay.\*

**Table 4.**
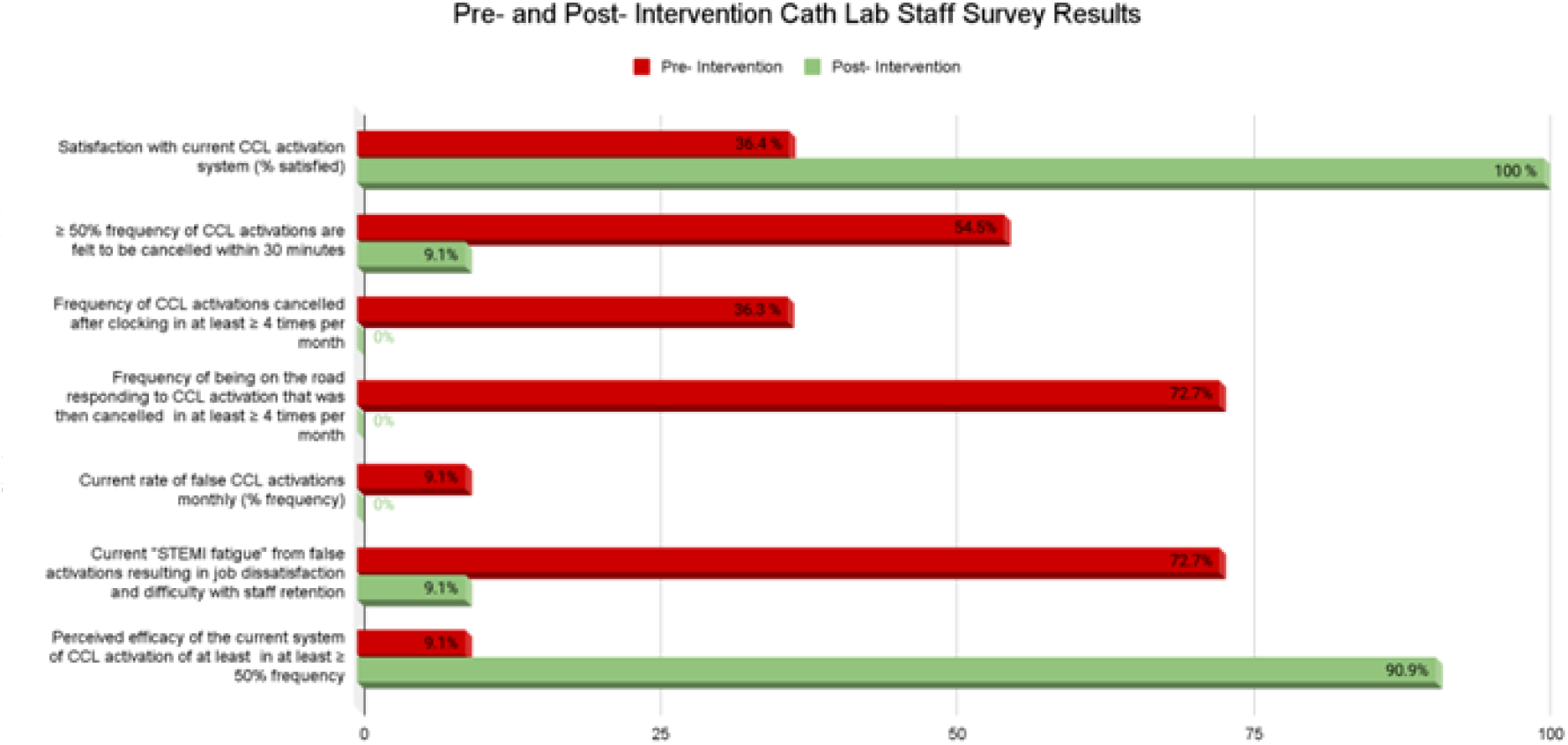
Cath Lab Staff Survey Results Pre--- and Post-Intervention.

## Discussion

In the final quarter of 2022, more than 60% of STEMI activations were canceled, with the majority of cases canceled following cardiologist evaluation. A new algorithm was implemented at our facility in early 2023, directing all out-of-hospital STEMI activations to the ED physician for review prior to activation of the STEMI alert and mobilization of CCL resources and staff. After a 30-day implementation plan, data collected on STEMI alerts and cancellations in the 2nd quarter of 2023 revealed a significant decrease in false activations to about 20%.

While false activations occur in almost every STEMI system, there is no national consensus or ACC/AHA recommendation regarding a safe or “acceptable” false-positive rate for CCL activation, which may reflect the diversity in STEMI systems of care and policy across the country. Available literature describes false positive rates ranging from 5% to 40%(6). A large 2007 prospective study at the Minneapolis Heart Institute by Larson et al found a false positive CCL activation of 14% for patients with suspected STEMI who underwent angiography without finding a culprit coronary arter. (7)Another study by Garvey et al examining CCL activation at fourteen primary angioplasty hospitals in North Carolina revealed an inappropriate STEMI activation rate of 15%. (8)

EMS agencies and first responders are a critical part of the STEMI system of care. Current protocol for suspected out-of-hospital STEMI involves initial medical screening and timely 12-lead ECG recording (9). The recommended methods of prehospital ECG interpretation involve trained paramedic evaluation, a computer algorithm, or transmission for physician interpretation. The accurate interpretation of this initial ECG in the context of symptom assessment is essential for triage and treatment decisions, including prehospital CCL activation, with previous studies showing early STEMI notification by EMS predicting shorter Door to Balloon (D2B) time and smaller infarct size. Early notification and prehospital CCL activation can also serve to reduce treatment times and eliminate the logistical burden caused by limited staffing, particularly on nights and weekends. A widely described drawback to pre-hospital activation of CCL is the frequent cancellation of alerts after EMS activation from the field. Previously evaluated reasons for cancellation in the literature include those that are patient-specific, such as goals of care, medical comorbidities, and contraindications to PCI. Other reasons involve systemic factors (ECG misinterpretation, reliance on computer readings, difficulties in prehospital ECG transmission).

Pre-intervention surveys conducted among CCL staff in 2022 revealed dissatisfaction with the existing activation system, with a significant proportion of personnel expressing concerns about a high incidence of false activations. “Alarm fatigue” is a well-documented phenomenon, particularly in intensive care settings, where clinicians and staff become desensitized to an overwhelming number of non-actionable alerts, potentially compromising patient safety. Inappropriate CCL activations incur significant costs, as on-call staff are required to respond, regardless of whether an emergent coronary angiography is performed. Additionally, CCL staff often leave their families and homes, sometimes at night, only to have activations canceled while en route, further contributing to dissatisfaction. This was reflected in the pre- and post-intervention surveys, where over 60% of false activations in 2022 negatively impacted staff morale and workplace satisfaction. After the implementation of our algorithm, the perceived effectiveness of the CCL activation system improved dramatically, rising from 9% to 90%.

Our action plan could be adapted and replicated at our institution to evaluate false positives in other multidisciplinary activation systems, such as Trauma and Stroke alerts, which are often initiated by EMS. While the algorithm was successfully integrated at our community hospital with input from all relevant stakeholders, larger tertiary care centers may face challenges in triaging patients solely based on ED evaluation, as opposed to immediate mobilization to the cath lab due to high patient volume. Other variables must also be considered from the moment EMS is contacted. ST-elevation myocardial infarctions (STEMI) represent a critical medical emergency with potentially high mortality rates, necessitating prompt recognition and intervention. In such scenarios, there is often a tendency to err on the side of caution due to patient complexity, physician preferences, and concerns about adverse patient or medico-legal outcomes. Although this approach is ethically reasonable, from a systems-level perspective, it can lead to financial burdens on both patients and hospitals, as well as contribute to caregiver burnout, as demonstrated in our pre-intervention surveys. While the American Heart Association has issued guidelines for STEMI care systems, local hospital authorities or government bodies ultimately determine triage, diagnosis, and treatment protocols at receiving facilities.

In Clay County, Florida, policies regarding STEMI care are determined at the hospital level. Our new CCL activation algorithm has removed the ability for EMS to trigger an out-of-hospital STEMI alert without prior review by an ED physician. Due to limited access to EMS data or CCL activations from other STEMI centers in our county, we were unable to assess the broader impact of our policy change. However, it would be valuable to evaluate EMS satisfaction with the new policy and explore whether there is a preference for transporting patients to other STEMI facilities in our area. In terms of ED physician responsibilities when activating a STEMI alert, clinicians at our facility frequently receive a single 12-lead ECG of variable quality from EMS, with limited opportunities for repeat assessments due to patient transport and support constraints. Improving data collection and transmission could enhance the STEMI care system at our institution.

In summary, our findings demonstrate that the modification of our pre-hospital STEMI activation policy led to a reduction in false activations of the CCL and PCI teams, accompanied by a subjective improvement in staff workplace satisfaction. The successful implementation of this initiative required collaboration and support from EMS, cardiologists, CCL staff, and ED physicians, along with all team members involved from the moment EMS is called.

While our study did not specifically examine changes in Door-to-Balloon (D2B) times, we observed no direct effect on D2B time following the implementation of the new STEMI activation algorithm.

## Conclusion

The multidisciplinary approach to the creation and implementation of the new STEMI activation algorithm resulted in a significant reduction in false activations, improved cath lab staff satisfaction, and better utilization of hospital resources.

## Data Availability

Not Applicable

## Declaration of interests

All authors have read and approved this manuscript for submission. We have each contributed substantially to the research and confirm no conflicts of interest to disclose. Furthermore, all authors agree to transfer copyright ownership to your journal upon publication.

Use of Artificial Intelligence: Grammarly and Quillbot were utilized to prepare this manuscript.

